# Risk of Polymyxin B-induced Acute Kidney Injury in Non-adjusted Dose Versus Adjusted Dose based on Renal Function: a retrospective cohort study

**DOI:** 10.1101/2020.11.23.20237479

**Authors:** Guanhao Zheng, Shenghui Zhou, Ning Du, Jiaqi Cai, Hao Bai, Juan He, Xiaolan Bian

**Affiliations:** Department of Pharmacy, Shenzhen Hospital, Southern Medical University, Shenzhen, Guangdong, China; Department of Pharmacy, Baiyin Central Hospital, Baiyin, Gansu, China; Department of Pharmacy, Qiqihar First Hospital, Qiqihar, Heilongjiang, China; Department of clinical laboratory, Kunshan Hospital Affiliated to Nanjing University of Chinese Medicine, Kunshan, Jiangsu, China; Department of pharmacy, Chongqing University Cancer Hospital, Chongqing, China; Department of Pharmacy, Ruijin Hospital Affiliated to Shanghai JiaoTong University School of Medicine, Shanghai, China

**Keywords:** Polymyxin B, Dose adjustment, Acute Kidney Injury, Nephrotoxicity, Renal function

## Abstract

**Background:** This study aimed to observe the difference in risk of polymyxin B-induced acute kidney injury with or without dose adjustment by patients’ renal function.

**Method:** A retrospective cohort analysis was carried out for patients who were treated with polymyxin B in Ruijin Hospital Affiliated to Shanghai Jiaotong University School of Medicine from November 2018 to October 2019. Patients were divided into adjusted dosage group and non-adjusted dosage group depended on dosage adjustment with renal function or not. A comparison of acute kidney injury incidence between the two groups was the primary outcome of this research. The secondary outcome included hospital length of stay, microbiological cure, clinical cure, and 30-day mortality.

**Result:** A total of 115 patients met the requirements of this study and were included in the analysis. Thirty-five patients were included in the non-adjusted dosage group and 80 in the adjusted dosage group. Patients from both groups had similar characteristics. The total daily dose of polymyxin B in the Non-adjusted dosage group was significantly higher than the adjusted dosage group (1.98 mg/kg/d vs 1.59 mg/kg/d, *P*=0.001). For the primary outcome of this research, no significant difference in the incidence of acute kidney injury was observed in these two groups (47.5% vs 37.14%, *P*=0.304), as well as the secondary outcomes, including hospital length of stay, microbiological cure, clinical cure, 30-day mortality.

**Conclusion:** Dosing adjustment renally could not lower the risk of polymyxin B-induced acute kidney injury significantly. A non-adjusted dosing strategy of polymyxin B is recommended when patients suffered from various levels of renal impairment.

## Background

Polymyxin B (PMB) is a kind of fundamental polypeptide antibiotics which derived from *Paenibacillus ploymyxa*[1-3]. It has been available to treat a series of bacterial infections since the 1950s due to its remarkable anti-microbial activity. However, PMB was gradually snubbed as an antimicrobial treatment option because of its severe adverse effect, particularly nephrotoxicity[4]. Owing to the lack of novel and effective antibiotics, PMB was attached great importance by clinicians again and regarded as the first-line treatment against multiple drug resistant (MDR) gram-negative bacteria in the last few decades[5, 6]. The nephrotoxicity associated with PMB especially has been of great concern since then.

It is reported that a high incidence of acute kidney injury (AKI) in patients receiving PMB ranges from 10 to 60%[7-13]. Dosage is one of the most crucial risk factors for PMB-induced AKI notably[9]. According to the research from Elias LS *et al*, a daily dosage greater than 200 mg was highly associated with the occurrence of severe renal impairment[14]. Dose adjustment by renal function is recommended for patients who receive PMB therapy in terms of drug information in the package insert of PMB.

However, the suggestion for optimal use of PMB is made that no dose adjustment should be applied when patients suffer various renal impairment extents because of the non-renal elimination characteristics of PMB, according to some pharmacokinetic/pharmacodynamic (PK/PD) studies[15-17]. Due to the inconsistency with the previous concept about PMB renally dosing adjustment and novel non-renal dose regimen, it is necessary to explore the efficacy and safety between these two dosing strategies.

The current study is primarily aimed to evaluate the incidence of AKI in patients treated with adjusted PMB dosage by renal function versus non-adjusted PMB dosage. The secondary objective of this research is about if dosage adjustment renally or not could significantly affect microbiological cure, clinical cure, and 30-day mortality.

## Methods

### Study population

This retrospective, single-center cohort study was carried out by the electronic medical record information system in Ruijin Hospital affiliated to the Medical School of Shanghai Jiao Tong University, China. Patients hospitalized from November 2018 to October 2019 and treated with PMB intravenously for MDR gram-negative infection were chosen as candidates for this study. Because of the observational, non-interventional design, the study was approved by Ruijin Hospital Institutional Review Board and performed by the ethical standards of the Declaration of Helsinki 1964 and its later amendments or comparable ethical standards. All treating and testing measure was informed consent by each patient. The eligibility criteria of participators were as follows: (1) Age between 18 and 85 years old; (2) Suspected or documented MDR gram-negative bacterial infection; (3) Treated by intravenous infusion of PMB for at least seven days. Patients with a creatinine clearance (CrCl) <10 mL/min calculated by Cockcroft-Gault formula[18] and/or suffering renal replacement therapy during the PMB treatment period were excluded in our research.

### Study objectives and variables

The primary objective of this research was to validate if there was a difference in the occurrence of AKI between PMB dosage adjusted by renal function or not. According to RIFLE criteria, AKI referred to serum creatinine (sCr) increasing for at least 1.5 times compared to baseline sCr on the 7^th^ day after initiation of PMB treatment, which was deeply classified as risk, injury, or failure based upon sCr increases of 1.5, 2.0, and 3.0 times baseline, respectively[19].

The secondary outcome of the current study was constituted by hospital length of stay (LOS), microbiological cure, clinical cure, 30-day mortality. Microbiological cure was defined as the elimination of pathogenic organisms during the PMB treatment period. Clinical cure was considered as the absence of clinical signs of infection after finishing the course of PMB therapy. 30-day mortality was identified as all-cause death within 30 days from initiation of PMB treatment.

### Polymyxin B dosing regimen

Researchers had separated eligible participators as the adjusted dosage group and the non-adjusted dosage group by dosing PMB with renal function or not. Each dose of PMB would have an infusing duration of no less than 60 minutes. As for the adjusted dosage group, patients with a CrCl greater than 80 mL/min received PMB 1.5-2.5 mg/kg per day in divided doses every 12 hours. Patients with CrCl between 30 to 80 mL/min would suffer lower dosage as 1.0-1.5 mg/kg every 24 hours, while patients with CrCl lower than 30 mL/min accepted 1.0-1.5 mg/kg every 2 to 3 days. All participators received the same dosing regimen as patients with CrCl >80 mL/min in the adjusted dosage group. No patients in either group received loading doses.

Participators’ demographics, comorbidity, characteristics at the initiation of PMB (age, sex, weight), past medical history (diabetes, cardiovascular disease, renal disease, hepatic disease), correlative laboratory results (baseline sCr and CrCl, sCr and CrCl on 7^th^ day of PMB therapy, sites of positive culture and isolated pathogens), PMB dosing regimen (loading dose, maintenance dose, and treatment duration), and concomitant nephrotoxic agents (aminoglycosides, vancomycin, loop diuretics, vasopressors, amphotericin B) during PMB therapy were recorded in detail from electronic medical record information system by researchers. Acute Physiology and Chronic Health Evaluation II (APACHE II) scores[20] were calculated and recorded when starting PMB therapy for all participators.

### Statistical analysis

All statistical analyses were conducted by SPSS (version 26, Chicago, IL, USA). The Mann-Whitney U test was utilized to compare continuous data, and the χ^2^ or Fisher exact test was performed for categorical data comparison.

## Results

### Patients’ demographics and characteristics

From November 2018 to October 2019, a total of 115 eligible patients fulfilled the inclusion and exclusion criteria in the present study, while 35 were involved in the non-adjusted dosage group and 80 in the adjusted dosage group.

All participators’ demographics and characteristics were listed in Table 1. Firstly, two groups had homologic characteristics with age, sex, and total body weight. Patients in the adjusted dose group owned non-significantly higher APACHE II scores and ICU admission rates. Moreover, a similar proportion was observed in each type of prior medical history between either group. There were also no differences in the concomitant use of all nephrotoxic agents between these two groups. As for baseline level and monitoring value after 7-day PMB treatment for both sCr and CrCl, no significant difference was found in the two groups.

**Table 1.** Patient demographics and characteristics

| Characteristic<br>(n=115) | Non-adjusted dosage<br>(n=35) | Adjusted dosage<br>(n=80) | P-Value |
| --- | --- | --- | --- |
| Age (years) | 60 (51-70) | 65 (51.25-73) | 0.235 |
| Sex |  |  |  |
| Male (%) | 25 (71.43) | 51 (63.75) | 0.424 |
| Female (%) | 10 (28.57) | 29 (36.25) |  |
| Total Body Weight (kg) | 65 (62.5-70) | 62 (55-71.38) | 0.913 |
| APACHE II score | 21 (17-28) | 23.5 (19-28.75) | 0.282 |
| ICU admission (%) | 21 (60) | 51 (63.75) | 0.702 |
| Prior medical history |  |  |  |
| Diabetes (%) | 6 (17.14) | 13 (17.5) | 0.906 |
| CVD (%) | 17 (48.57) | 41 (51.25) | 0.792 |
| Renal Disease (%) | 14 (40) | 34 (42.5) | 0.802 |
| Hepatic Disease (%) | 14 (40) | 23 (28.75) | 0.235 |
| Serum creatine concentration (μmol/L) |  |  |  |
| Baseline | 75 (43-105) | 69 (52-92) | 0.775 |
| Day-7 PMB treatment | 99 (79-167) | 117 (73-176) | 0.889 |
| Creatine clearance (mL/min) |  |  |  |
| Baseline | 80.93 (60.57-127.43) | 85.24 (60.68-119.75) | 0.998 |
| Day-7 PMB treatment | 57.79 (34.48-74.09) | 58.55 (32.36-83.22) | 0.853 |
| Concomitant nephrotoxic agents |  |  |  |
| Aminoglycosides (%) | 2 (5.71) | 2 (2.5) | 0.387 |
| Vancomycin (%) | 3 (8.57) | 8 (10) | 0.811 |
| Loop diuretics (%) | 11 (31.43) | 24 (30) | 0.878 |
| Vasopressors (%) | 10 (28.57) | 29 (36.25) | 0.424 |
| Amphotericin B (%) | 0 (0) | 2 (2.5) | 0.345 |
| Sites of positive culture |  |  |  |
| Blood (%) | 19 (54.29) | 30 (37.5) | 0.094 |
| Respiratory tract (%) | 21 (60) | 61 (76.25) | 0.076 |
| Urine (%) | 2 (5.71) | 3 (3.75) | 1.000 |
| Wound (%) | 5 (14.29) | 8 (10) | 0.728 |
| Other (%) | 3 (8.57) | 10 (12.5) | 0.770 |
| Isolated pathogen |  |  |  |
| *Klebsiella pneumoniae* (%) | 23 (65.71) | 64 (80) | 0.100 |
| *Pseudomonas aeruginosa* (%) | 8 (22.86) | 21 (26.25) | 0.700 |
| *Acinetobacter baumannii* (%) | 16 (45.71) | 32 (40) | 0.567 |
| Other organisms (%) | 1 (2.86) | 3 (3.75) | 1.000 |

Data are median (interquartile range, IQR) values or no. (%) of patients.
APACHE = Acute Physiology and Chronic Health Evaluation.
CVD = Cardiovascular Diseases.

The most common site of positive culture in the adjusted dosage group was the respiratory tract, which had an insignificantly higher rate than the non-adjusted dosage group (76.25% vs 60%, *P*=0.076). Blood was another crucial site of infections with significantly different detection rates between these two groups, while 54.29% in the non-adjusted dosage group and 37.5% in the adjusted dosage group (*P*=0.094). More *Klebsiella pneumoniae* (80% vs 65.71%, *P*=0.100) were isolated in the adjusted dosage group than non-adjusted dosage group as dominant isolated pathogens in this research. Both groups showed no significant difference in the isolated rate of *Pseudomonas aeruginosa* (22.86% vs 26.25%, *P*=0.700) and *Acinetobacter baumannii* (45.71% vs 40%, *P*=0.567), respectively.

### Polymyxin B dosing details

The PMB dosing details were exhibited in Table 2. The average PMB therapy duration showed no significant difference between each group (9d vs 12.5d, *P*=0.126). The total daily dose in mg/kg/d for the non-adjusted dosage group was significantly higher than the adjusted dosage group (1.98 mg/kg/d vs 1.59 mg/kg/d, *P*=0.001).

**Table 2.**
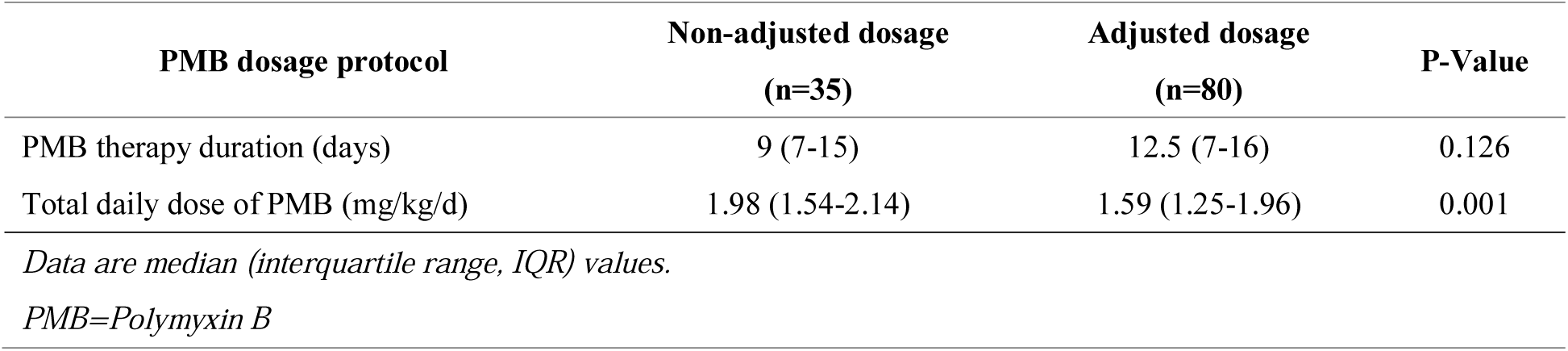
PMB dosage details

### Primary and secondary outcomes

When it comes to the primary outcome of current research (Table 3), patients with adjusted PMB dosage had a higher but statistically insignificant incidence of AKI than those who received a non-adjusted dosage of PMB (47.5% vs 37.14%, *P*=0.304). Patients who suffered from AKI had a similar distribution of various injury levels, whether with or without PMB dosage adjustment. Secondary outcome (Table 3) indicated that better microbiological cure (37.14% vs 31.25%, *P*=0.536) and higher clinical cure rate (45.71% vs 33.75%, *P*=0.222) existed in the non-adjusted dosage group, as well as lower 30-day mortality (28.57% vs 38.75%, *P*=0.294), although a significant difference was not found for these parameters. Days of LOS did not differ significantly in these two groups (55d vs 53.5d, *P*=0.513).

**Table 3.** Primary and secondary outcomes

| Outcome | Non-adjusted dosage | Adjusted dosage | P-Value |
| --- | --- | --- | --- |

|  | (n=35) | (n=80) |  |
| --- | --- | --- | --- |
| **Primary Outcomes** |  |  |  |
| AKI (%) | 13 (37.14) | 38 (47.5) | 0.304 |
| Risk/KDIGO stage 1 (%) | 5 (38.46) | 15 (39.47) | 0.561 |
| Injury/KDIGO stage 2 (%) | 4 (30.77) | 11 (28.95) | 0.734 |
| Failure/KDIGO stage 3 (%) | 4 (30.77) | 12 (31.58) | 0.611 |
| **Secondary Outcomes** |  |  |  |
| LOS (days) | 55 (26-91) | 53.5 (40.25-89.75) | 0.513 |
| Microbiological cure (%) | 13 (37.14) | 25 (31.25) | 0.536 |
| Clinical cure (%) | 16 (45.71) | 27 (33.75) | 0.222 |
| 30-day mortality (%) | 10 (28.57) | 31 (38.75) | 0.294 |

*Data are median (interquartile range, IQR) values or no. (%) of patients.*
*KDIGO=Kidney Disease: Improving Global Outcomes*
*LOS=Length of Stay*

## Discussion

Nowadays, MDR gram-negative bacteria show a severe threat to patients’ anti-infective treatment, leading to high morbidity and mortality, especially for critically ill patients[21]. Carbapenem-Resistant Enterobacteriaceae (CRE) refers to a kind of gram-negative bacteria with multiple drug resistance, which could be contagious and even fatal. The rapid spread of CRE has become a global health crisis over the last decade. It is challenging to deal with this emerging bacterial threat all over the world[22-24].

Besides other effective antibiotics like tigecycline and ceftazidime/avibactam, PMB plays an essential role as a first-line agent with its reliable capability against CRE[25-29]. It is urgent to determine the appropriate PMB dosing strategy for optimal use due to its dose-related nephrotoxicity. Since PMB is a kind of concentration-dependent antibiotic, higher exposure of PMB is correlated with better bacterial killing capability, which means a subtherapeutic dose of PMB could give rise to microbiological treatment failure and bacterial resistance[30-32]. In other words, dosage is a vital factor on the efficacy and safety of PMB clinical use.

Hence, it is reasonable to evaluate clinical efficacy and safety of PMB dosing adjustment with or without kidney function. The dominating purpose of our research is to explore if adjusting the PMB dose regimen by renal function could lower the incidence of AKI. Assessment of LOS, microbiological cure, clinical cure, and 30-day mortality are made to compare these two different dosing regimens.

In the current study, dosing adjustment by patients’ renal function even resulted in a non-significant higher incidence of AKI than the non-adjusted dosing strategy. Since patients in each group had similar characteristics and demographics, renally dosing adjustment was not an effective dosing strategy to reduce the risk of PMB-induced AKI based upon our research result.

However, it was controversial that patients with renally dosing adjustment had a higher risk of AKI with a significantly lower total daily dose of PMB, which was discrepant with the research result of Rigatto *et al* that total dose of PMB was highly positive correlated with the incidence of AKI[9]. A reasonable explanation of this controversy is that severe infection is a prime cause of AKI for critical ill patients, especially for patients with sepsis. There was a significantly lower PMB dose for patients who suffered from various levels of renal impairment with renally dosing adjustment. An insufficient dose was employed when patients received PMB therapy against infection accompanied by renal dosing adjustment, causing the failure of bacteria elimination, exacerbation of persistent infection, even emergence of sepsis or multi-organ dysfunction, including kidney impairment would develop AKI eventually[33]. The pathogenesis of AKI in severe infection or sepsis is highly related to renal hypoperfusion derived from the toxic effects of inflammation and impaired microcirculation[34]. A single-center, retrospective study with 54 patients was implemented by Maniara *et al* also obtained a similar result that subtherapeutic doses from renal adjustment could lead to unresolved infections and a higher incidence of AKI[35]. In consequence, renal dosing adjustment of PMB is unnecessary for patients with kidney function impairment.

Besides the risk of AKI, a comparison of clinical efficacy between these two dosing strategies was also carried out. Patients in our study who received PMB treatment with renal adjustment had lower microbiological cure rate and clinical cure rate and higher 30-day mortality. This phenomenon was attributed to the subtherapeutic dose of PMB, which could result in uncontrolled infection and sepsis, increasing infection-induced mortality. Peyko *et al* found that patients who received renally adjusted PMB doses owned higher LOS and mortality due to PMB underdose against infection[36]. Research from Maniara *et al* showed that renal dose adjustment of PMB did not affect microbiological cure, clinical cure, or 30-day mortality[35]. In this aspect, dosing without renal adjustment is a proper PMB dosing strategy to increase the probability of meeting the therapeutic target and get a better curative rate ultimately.

There are some limitations to this study. Firstly, this was a retrospective, observational study with an inadequate size of participators. Further investigation should be performed as a Randomized Controlled Trial (RCT) or other prospective researches. Secondly, sCr level evaluation in terms of Cockcroft-Gault Equation could be likely to overestimate renal function for the elders, leading to falsely low circulating creatinine or normal sCr with actually decreased renal function[37]. Therefore, evaluating AKI risks could not be more specific in our studies due to patients’ relatively high average age. Besides, measurement of AKI incidence without recording specific AKI onset time and daily sCr monitoring could lead to inexact results for patients who suffered AKI previously and recovered before day-7 measurement. Finally, some studies have elucidated that an average steady-state plasma concentration of 2 mg/L was the appropriate PK/PD therapeutic target of PMB since increasing incidence and severity of AKI were seen in higher PMB concentration than this[38]. It is valid to find out the relationship between various dosing strategies and the incidence of AKI through therapeutic drug monitoring (TDM) of PMB in our next researches.

## Conclusion

In conclusion, the PMB dose adjustment by renal function could barely affect AKI incidence based on the current study compared to the non-adjusted dosing strategy. There was also no significant difference between these two dosing strategies for all secondary outcomes, including LOS, microbiological cure, clinical cure, or 30-day mortality. Our findings are consistent with prior similar clinical studies about dose adjustment renally or not. Due to the dose-related bactericidal activity of PMB, the non-adjusted dosing strategy is recommended for patients with renal impairment while it owns a higher probability of eradicating susceptible pathogens without increasing the risk of PMB-induced AKI. Further research is needed to re-evaluate the outcomes described above to provide adequate evidence supporting the non-adjusted dosing regimen of PMB.

## Data Availability

All data and material used and/or analyzed during the current study are available from the corresponding author on reasonable request.

## Abbreviations

PMB: polymyxin B
MDR: multiple drug resistant
AKI: acute kidney injury
PK/PD: pharmacokinetic/pharmacodynamic
CrCl: creatinine clearance
sCr: serum creatinine
LOS: length of stay
APACHE II: Acute Physiology and Chronic Health Evaluation II
CRE: Carbapenem-Resistant Enterobacteriaceae
RCT: Randomized Controlled Trial
TDM: therapeutic drug monitoring.

## Declarations

### Ethics approval and informed consent to participate

This study was approved by Ruijin Hospital Institutional Review Board and has been performed in accordance with the ethical standards laid down in “Declaration of Helsinki 1964” and its later amendments or comparable ethical standards. Written informed consent was obtained from individual or guardian participants.

### Consent to publication

Not applicable.

## Conflicts of interest

All authors declared no conflict of interest in this study or the findings specified in this paper.

## Funding

The authors received no specific funding for this work.

## Authors’ contributions

GZ, JH and XB conceived and designed this study. GZ, SZ, ND, JC, HB and XB collected the information in the case, and contributed to the acquisition, analysis, and interpretation of the data. GZ, JH and XB wrote and revised the manuscript. All authors read and approved the final manuscript.

### Acknowledgements

The authors thank the staff of the pharmacy and EICU Department of Ruijin Hospital affiliated to Medical School of Shanghai Jiao Tong University for their facilities and collaboration.

## Authors’ information

1: Department of Pharmacy, Shenzhen Hospital, Southern Medical University, Shenzhen, Guangdong, China;

2: Department of Pharmacy, Baiyin Central Hospital, Baiyin, Gansu, China;

3: Department of Pharmacy, Qiqihar First Hospital, Qiqihar, Heilongjiang, China;

4: Department of clinical laboratory, Kunshan Hospital Affiliated to Nanjing University of Chinese Medicine, Kunshan, Jiangsu, China.

5: Department of pharmacy, Chongqing University Cancer Hospital, Chongqing, China;

6: Department of Pharmacy, Ruijin Hospital Affiliated to Shanghai JiaoTong University School of Medicine, Shanghai, China;

7: Department of Pharmacy, Ruijin Hospital Affiliated to Shanghai JiaoTong University School of Medicine, Shanghai, China.

## Notes

### Competing Interest Statement

The authors have declared no competing interest.

### Funding Statement

The authors received no specific funding for this paper.

### Author Declarations

This study was approved by Ruijin Hospital Institutional Review Board and has been performed in accordance with the ethical standards laid down in Declaration of Helsinki 1964 and its later amendments or comparable ethical standards.

